# Comparing outcomes following a first episode of psychosis in autistic and non-autistic people: a clinical retrospective cohort study

**DOI:** 10.64898/2026.06.01.26354576

**Authors:** John H Ward, Jonathan Lewis, Elizabeth M Weir, Tamsin J Ford, Rudolf N Cardinal

## Abstract

**Background:** There is growing evidence to suggest a clinically significant overlap between autism spectrum conditions and psychotic disorders. Preliminary evidence suggest that autism diagnoses and autistic traits are associated with poorer outcomes following a first episode of psychosis.

**Methods:** This study used data from the Cambridgeshire and Peterborough National Health Service Foundation Trust (CPFT) Research Database to examine clinical outcomes in autistic and non-autistic people following a first episode of psychosis. We describe patterns of community and inpatient service use, using descriptive statistics, Cox regression, binomial logistic regression, and negative binomial regression.

**Results:** Data from 224 autistic and 7185 non-autistic people with psychosis were analysed. Autism was associated with greater community service use (use of mental health emergency lines, mental health detentions by police), as well as greater likelihood of psychiatric hospital admission (adjusted hazard ratio 1.42, 95% confidence interval 1.08–1.88, p<0.05) and longer inpatient stays (median 112 versus 49 days, p<0.0001). Learning disability played a significant role in the utilisation of community and inpatient services, with lower rates of community service use but longer inpatient admissions.

**Conclusions:** This study indicates a differing pattern of service use between autistic and non-autistic people following psychosis that warrants further research into how best to support autistic people with psychosis.

**Lay Summary:** Autistic people are at greater risk of developing psychosis. This study aimed to compare the outcomes and mental health service use of autistic people with and without psychosis. This study found that autistic people with psychosis were at greater risk of hospitalisation following a psychosis diagnosis. We found that autistic people were more likely to have used emergency mental health services relative to non-autistic people. We also found that people with a learning disability in addition to autism and psychosis used services in different ways.

---

There is growing evidence demonstrating an association between autism and psychotic disorders (also known as schizophrenia spectrum disorders, henceforth ‘psychosis’) (Ahi Üstün, Yazıcı, İlhan, & Saka, 2025; Jeong et al., 2024; Kincaid, Doris, Shannon, & Mulholland, 2017; Vaquerizo-Serrano, Salazar de Pablo, Singh, & Santosh, 2022). In England, approximately 0.82% of adults have diagnosed autism (with a further 0.77–2.12% suspected undiagnosed) (O’Nions et al., 2023), whilst approximately 0.4% of adults have a psychotic disorder (Morris, Hill, Brugha, & McManus, 2025).

Meta-analytic evidence estimates a 3–4-fold increased risk for developing psychosis in autistic people (Z. Zheng, Zheng, & Zou, 2018). Crucially, evidence suggests that these autistic traits in psychosis are stable through treatment, and thus not a matter of converging autistic/psychosis traits, although overlap in clinical features is recognised (Chisholm et al., 2019).

Primary psychotic disorders are considered severe mental illnesses, a term recognising potential chronicity and significant impacts on social/occupational functioning, health, quality of life, and mortality (Correll et al., 2022; de Winter et al., 2022, 2023; Moreno et al., 2013; Peritogiannis, Gogou, & Samakouri, 2020; Zumstein & Riese, 2020). These impacts are partially mitigated by prompt treatment to reduce the duration of untreated psychosis (DUP), which in England is delivered via Early Intervention in Psychosis (EIP) services (de Pablo et al., 2024; Fern, Regan, & Lucas-Motley, 2025; Puntis et al., 2020; White et al., 2009). EIP services are commissioned to provide care for up three years after a first episode of psychosis (FEP) and involve a broad range of professional expertise, including psychiatrists, psychologists, family therapists, care coordinators, and support workers (Fern et al., 2025; National Institute for Health and Care Excellence, 2015).

There is preliminary evidence that autistic people have poorer health and social outcomes following FEP. A study of 180 Singaporean EIP patients found that higher Autistic Spectrum Quotient (AQ-10) scores were associated with poorer improvements in the Positive and Negative Symptoms Scale (PANSS, for schizophrenia) and poorer global functioning at follow-up (S. Zheng et al., 2021). Further studies have found that neurodivergent people spend longer in mental health hospital following FEP, and this has previously been linked to DUP, a recognised determinant of long-term outcomes (Ajnakina et al., 2020; de Pablo et al., 2024; Oduola, Craig, Iacoponi, Macdonald, & Morgan, 2024). Unfotunately, autistic adolescents with psychosis have higher rates of treatment failure (Downs et al., 2017; Nikolić, Sculthorpe, Stock, Stevens, & Eccles, 2025).

In a broader context, autistic people have higher rates of physical and mental health morbidity (Martini et al., 2022; John H. Ward, Weir, Allison, & Baron-Cohen, 2023; Weir, Allison, Warrier, & Baron-Cohen, 2021), poorer experiences of healthcare (NHS Digital, 2025; Radev, Freeth, & Thompson, 2024; Vogan, Lake, Tint, Weiss, & Lunsky, 2017), and reduced life expectancy (O’Nions et al., 2024).

Therefore, the aim of this study was to characterise the outcomes following first-episode psychosis in autistic and non-autistic people, supported by descriptive and inferential quantitative analyses of anonymised clinical records.

## Methods

### Study Design

This study was a retrospective cohort study, using data collected in routine clinical practice from patients under the care of Cambridgeshire and Peterborough NHS Foundation Trust (CPFT), a mental health and community services Trust in the East of England affiliated with the University of Cambridge (Cardinal, 2017). The research database collects clinician data on an opt-out basis. De-identified records from 2013 onwards were manipulated and extracted for analysis using Microsoft SQL Server. This study was ethically approved (NHS Research Ethics approval for the CPFT Research Database, 22/EE/0264; CPFT Research Database Oversight Committee study reference Z0065).

### Participants

The inclusion criterion for this study was anyone within the CPFT Research Database who had a psychotic disorder diagnosis of interest (**Table 1**) recorded within their clinical record at the time of data extraction (4 March 2026). Where someone had multiple psychotic disorder diagnoses (e.g. unspecified non-organic psychosis, followed by schizophrenia), the first diagnosis was taken as the point of entry into the study.

**Table 1:** Relevant diagnoses (International Classification of Disease [ICD]-10 codes, ‘x’ as single-character wildcard)

| Psychosis Diagnoses (ICD-10) |
| --- |
| Drug/substance-induced psychoses (F10–F19, F1x.5) |
| Schizophrenia (F20) |
| Schizotypal disorder (F21) |
| Persistent delusional disorder (F22) |
| Acute and transient psychotic disorder (F23) |
| Schizoaffective disorder (F25) |
| Other non-organic psychotic disorders (F28) |
| Unspecified non-organic psychosis (F29) |
| Hypomania/mania (F30), bipolar affective disorder (F31) |
| Depression with psychotic features (psychotic depression) (F32.3, F33.3) |

Autistic people were identified in two ways. Firstly, they were identified via a recorded diagnosis of autism in their clinical record as described above (ICD-10 code F84). Given the long waiting lists for neurodevelopmental services, we also included people in our autistic group who were on a waiting list for an autism service (either child or adult). We present results from the sensitivity analyses of people with diagnosed autism only in the **Supplementary Files**.

### Data extraction

#### Covariates

Where available, relevant covariates (gender, ethnicity, Index of Multiple Deprivation [IMD] decile) were extracted for each person. The platform for clinical records changed within CPFT in 2020–2021 (from Servelec RiO to TPP SystmOne), after which the former RiO database was no longer updated. Where conflict arose between datasets, SystmOne data was chosen in preference, as the most recent dataset.

#### Community/Inpatient Service Use Information

Data on relevant community services were extracted. These were; early intervention in psychosis services, urgent mental health phone lines (nationally NHS 111 option 2, known locally as the ‘First Response Service’), Section 136 utilisation (mental health act (MHA) legislation for the police to remove someone with a suspected mental disorder from the public to a place of safety) and crisis and resolution home treatment team (CRHTT, also known as the crisis team).

Data on inpatient admissions were also extracted, including dates and uses of MHA frameworks. Due to limitations in the data, out-of-area admission ((i.e. admissions of patients outside of CPFT) were captured as a binary variable (lifetime out-of-area admission yes/no).

Full details of how community, inpatient and out-of-area data were extracted are given in **Supplementary Files.**

#### Medication Usage

Medications were identified in patient records using natural language processing (NLP, using General Architecture for Text Engineering [GATE] software) of free text data, as per methods previously used on this dataset (Cardinal, Savulich, Mann, & Fernández-Egea, 2015). This extraction had occurred fully only within the RiO dataset, and therefore these analyses are based on a smaller subset of the population (6473 non-autistic and 207 autistic participants). Antipsychotic prescriptions were selected from a prespecified list of antipsychotics (based on a combination of the British National Formulary and the local NHS Trust formulary), with an algorithm to determine whether these were likely to be long-acting injectable prescriptions (LAIs, also known as ‘depots’; see **Supplementary Files**).

### Data Cleaning

#### Psychosis Diagnoses

For the purposes of statistical disclosure control and data analysis, some diagnoses were aggregated. Bipolar affective disorder and ‘manic episode’ were aggregated into ‘bipolar affective disorder (including mania)’; acute and transient psychotic disorders and other non-organic psychotic disorders were also merged into ‘acute and transient psychotic disorders (including other nonorganic psychotic disorders)’.

#### Ethnicity

Ethnicity was aggregated based on statistical disclosure and interpretability into six broad categories; White, Black, Asian, Mixed, Other and Missing (in line with UK Census reporting) (Office for National Statistics, 2022).

### Data Analysis

Data were first examined using descriptive methods, followed by basic transformations (means, standard deviations, medians, interquartile ranges [IQRs]) and basic parametric/non-parametric statistical tests (*t* test, Wilcoxon signed rank test, chi-squared tests).

Cox proportional hazard ratios were calculated on the time-to-event data from time of psychosis diagnosis to first admission either informally (‘voluntary’ admission) or involuntary (detained) admission. These models were calculated firstly unadjusted, then adjusted by basic covariates (age at psychosis diagnosis, ethnicity, IMD decile, gender), before finally adjusting for recorded learning disability (LD) diagnosis (see below).

To examine data on the durations of care under psychiatric intensive care units or CRHTT, negative binomial logistic regression models were used, adjusted as for the Cox regression models.

Finally, negative binomial regression was used to examine the relationship between autism and the total lifetime number of antipsychotics trialled, whilst binomial logistic regression was used to predict the likelihood of ever receiving a LAI antipsychotic.

#### Exploratory analysis

Data on learning disability (LD) (derived from ICD-10 codes F70–79) were extracted from patients’ clinical records and then used to further subdivide groups, given a hypothesis that groups with autism and LD may be characteristically different from those with just autism. Data were split into four categories: control (neither autism nor LD), autism only, LD only, autism and LD. Data were first analysed using a basic four-way comparison (via analysis of variance [ANOVA], Kruskal–Wallis, or chi-squared tests). Comparisons that yielded significant results were analysed further using a regression model with autism, LD (binary), and interaction (autism × LD) terms. These were performed unadjusted, and then adjusted for diagnosis, age at psychosis diagnosis, ethnicity, gender, and IMD decile.

#### Sensitivity analysis

As the autism group comprised both diagnosed autism and people awaiting diagnosis, we performed a sensitivity analysis that limited to only those people with diagnosed autism (those on waiting lists without diagnosis were removed from the analysis entirely).

### Patient and Public Involvement and Engagement (PPIE)

This work was supported by patient, public, and clinician involvement and engagement, to understand the importance of this topic for autistic people with psychosis, their families, and clinicians, as well as their priorities for research. PPIE sessions took the form of 1-hour conversations with participants, aided by a few pre-devised promptingquestions. These conversations happened just prior to analysing the data. Part of the engagement discussions focussed explicitly on how autistic people, families and clinicians measure progress in recovery. We received strong feedback that this was an important and under-appreciated area, particularly in reference to the experiences of psychiatric care among autistic people, delayed/missed autism diagnosis, and long-term outcomes following an autism diagnosis.

### Pre-print registration

Prior to publication, this manuscript was uploaded to medRxiv; a widely recognised preprint server (Ward, Lewis, Weir, Ford, & Cardinal, 2026).

## Results

This study explored outcomes for 7,409 people with psychosis, including 224 people with diagnosed or suspected autism. Of those 224 people, 185 had a confirmed diagnosis of autism, whilst 39 were awaiting diagnostic assessment for autism.

The number of males in the autistic group was significantly higher than in the non-autistic group (**Table 2)**. The missingness of the ethnicity data was also much lower in the autistic group than the non-autistic group (12.1% versus 23.7%). The missingness of gender data was <1% for both the autistic and non-autistic groups.

**Table 2:**
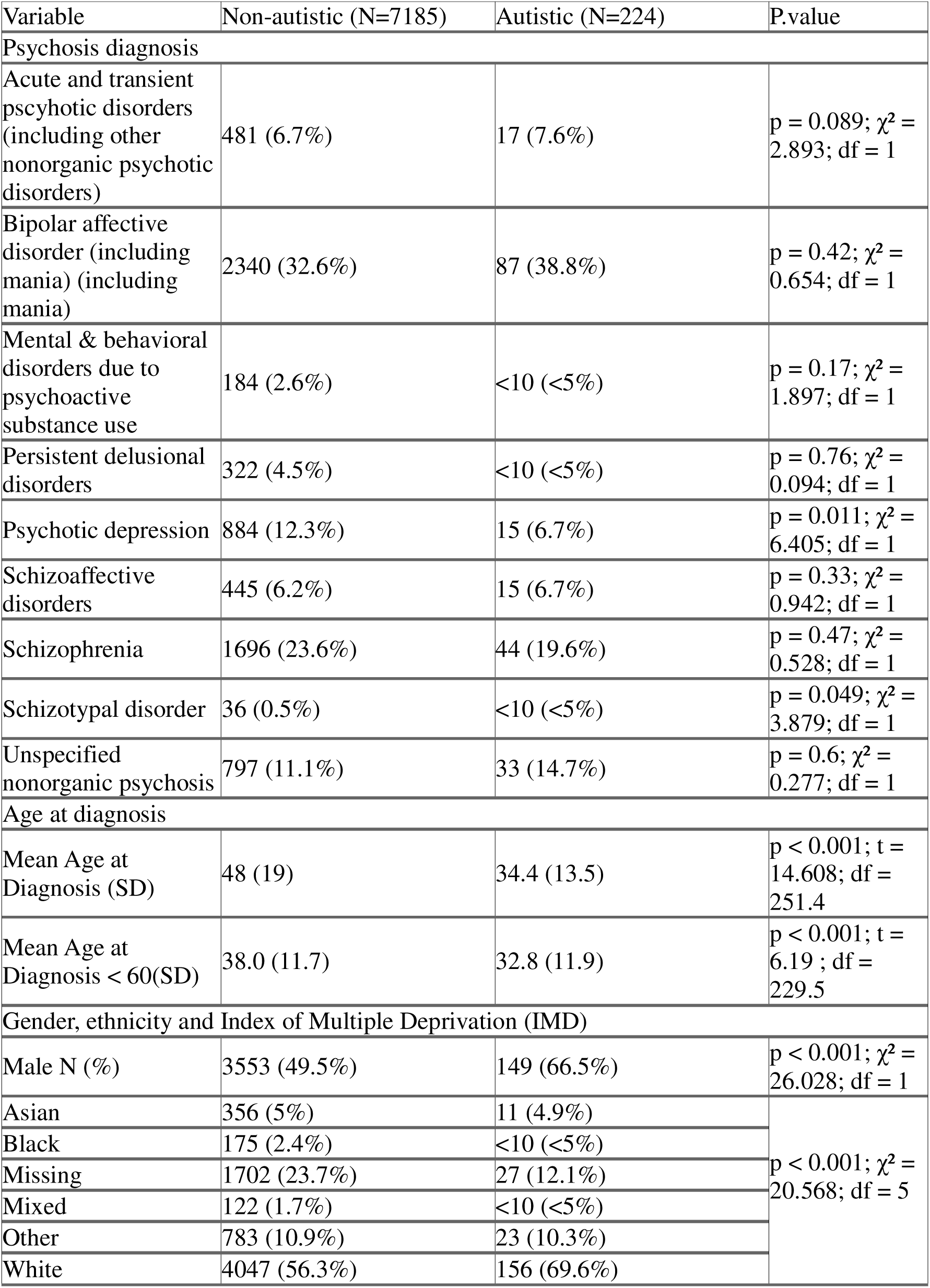
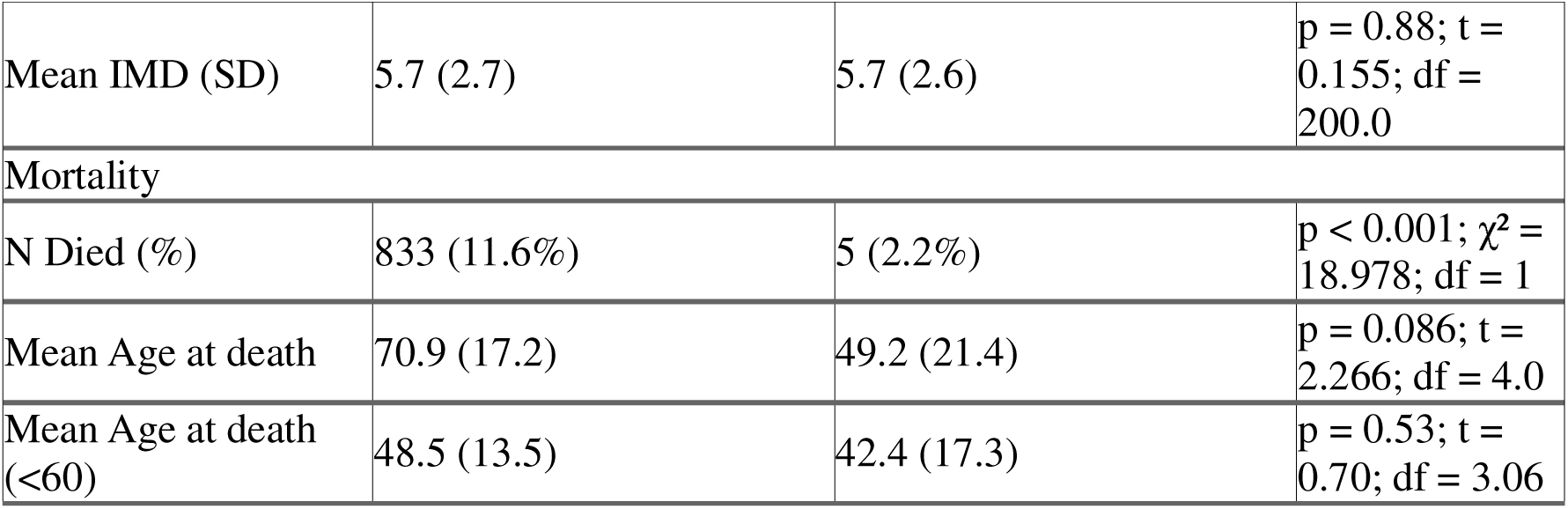
Demographic information for non-autistic and autistic people with psychotic disorders. IMD, Index of Multiple Deprivation.

Across both groups, the most common psychiatric diagnosis was bipolar disorder, followed by schizophrenia (F20) and unspecified non-organic psychosis (F29) (**Table 2**). The rates of conversion from F29 to F20 were non-significant between groups (10/39 autistic, 115/791 non-autistic, χ^2^ = 3.38, df=1, p>0.05). Autistic people were diagnosed with psychosis at a younger age on average, with a large number of the non-autistic patients (29.1%) diagnosed after the age of 60 (compared with 4.9% of the autistic group) (**Table 2, Table S1 (see Supplementary Files**)).

A small proportion of patients had died at the point of data extraction (<5% of the autistic group, 11.6% of the non-autistic group). There was no sigificant difference in the mean age at death (49.2 years autistic versus 70.9 years non-autistic, t=2.26, df=4.03, p>0.05), and this was also true when excluding people diagnosed with psychosis after the age of 60 (**Table 2**).

Our data also demonstrated that autism and psychosis diagnoses were often made at similar times, but with a wide range (e.g. some autism diagnoses made 20–30 years after diagnosis of psychosis) (**Figure S1** (**see Supplementary Files))**.

### Community Service Use

There were no significant differences between groups in rates of referral to EIP or number of referrals rejected as ‘inappropriate’. The number of appointment contacts and completion of treatment (i.e. three years under the care of EIP, as intended) was also similar between groups (56.6% autistic versus 44.6% non-autistic).

Patients in the non-autistic group were more likely to have seen a liaison (consultation/liaison) psychiatry service (LPS) than the non-autistic group (χ² = 6.645, df = 1, p<0.01), although the median number of LPS contacts per patient did not differ (**Table 3**).

**Table 3:** Service use data for people with psychotic illnesses, by group. EIP=Early Intervention in Psychosis Services, LPS=Liaison Psychiatry Services, FRS=First Response Service, CRHTT=Crisis Resolution and Home Treatment Team. PICU= psychiatric intensive care unit. MHA=Mental Health Act (i.e. detained).

| Variable | Non-autistic (N=7185) | Autistic (N=224) | P.Value |
| --- | --- | --- | --- |
| Referred to EIP (%) | 1268 (17.6%) | 46 (20.5%) | $p = 0.27; \chi^2 = 1.242; df = 1$ |
| Inappropriate Referral (%) | 198 (15.6%) | 10 (21.7%) | $p = 0.26; \chi^2 = 1.249; df = 1$ |
| Treatment Completed (%) | 565(44.6%) | 26(56.5%) | $p = 0.11; \chi^2 = 2.567; df = 1$ |
| Mean Contact (SD) | 13 (16.3) | 14.5 (18.9) | $p = 0.33; t = -0.978; df = 156.6$ |
| Seen by LPS (%) | 3418 (47.6%) | 87 (38.8%) | $p = 0.0099; \chi^2 = 6.645; df = 1$ |
| Median LPS Contact (IQR) | 2 (1-3) | 2 (1-3.5) | $p = 0.5; W = 142784$ |
| Lifetime S136 (%) | 349 (4.9%) | 20 (8.9%) | $p = 0.0058; \chi^2 = 7.608; df = 1$ |
| S136 Count Median (IQR) | 2 (1-4) | 2 (1-3.2) | $p = 0.99; W = 3497$ |
| Lifetime FRS (%) | 2655 (37%) | 95 (42.4%) | $p = 0.096; \chi^2 = 2.773; df = 1$ |
| FRS Count Median (IQR) | 2 (1-16) | 3 (1-21) | $p = 0.0088; W = 106927$ |
| Lifetime CRHTT (%) | 4378 (60.9%) | 100 (44.6%) | $p < 0.001; \chi^2 = 24.108; df = 1$ |
| Median Number of CRHTT Referrals (IQR) | 2 (1-5) | 2.5 (1-6) | $p = 0.15; W = 200751$ |
| Proportion of males admitted to PICU (%) | 354 (10%) | 14 (9.4%) | $p = 0.82; \chi^2 = 0.051; df = 1$ |
| Median PICU Length of Stay (IQR) | 25 (12-49) | 35 (19.2-46.8) | $p = 0.42; W = 2168$ |
| Admitted Informally N(%) | 2760 (38.4%) | 93 (41.5%) | $p = 0.35; \chi^2 = 0.884; df = 1$ |
| Median Admission Duration (Informal Admissions) | 31 (12-81) | 52 (13-146) | $p = 0.018; W = 109810$ |
| Admitted Under MHA N(%) | 1865 (26%) | 69 (30.8%) | $p = 0.1; \chi^2 = 2.645; df = 1$ |
| Median Admission Duration (MHA Admissions) | 55 (25-122) | 74 (36-180) | $p = 0.032; W = 54592$ |
| Admitted Ever N (%) | 3752 (52.2%) | 118 (52.7%) | $p = 0.89; \chi^2 = 0.018; df = 1$ |
| Median Admission Duration (MHA and | 49 (19-121) | 112 (24.8-213.2) | $p < 0.001; W = 168845$ |
| Informal) |  |  |  |
| Admitted Out of Area<br>N(%) | 257 (3.6%) | 15 (6.7%) | p = 0.014; $\chi^2 =$<br>5.978; df = 1 |

Autistic people were more likely to have been subject to a Section 136 police detention than non-autistic people (8.9% vs 4.9%, p<0.01). However, there was no difference in the median number of S136 contacts. Autistic people were also more likely to have utilised the telephone crisis First Response Service (45.7% versus 36.8%), but this different was non-statistically significant. However, autistic patients on average had a greater number of FRS contacts (median 3 [IQR 1–21] autistic, versus 2 [IQR 1–16] non-autistic, p<0.01).

Interestingly, autistic people were less likely to have ever been referred to a crisis team (44.6% vs 60.9%, p<0.0001), but with no difference in number of referrals per patient.

### Inpatient Service Use

When combining both voluntary and involuntary admissions, autistic people were more likely to be admitted at any point in time in the study (adjusted hazard ratio (aOR) 1.42, 95% confidence interval (95% CI) 1.08-1.88, p<0.05), including when adjusting for learning disability **(Table 3)**. When subdividing by admission type, this effect only held for voluntary admissions and disappeared when adjusting for learning disability.

Analysis of psychiatric intensive care unit (PICU) admissions only examines males, as CPFT only provides male PICU beds (**Table 2**). There was not a significant difference in the proportion of males admitted to psychiatric intensive care (PICU) between autistic and non-autistic males (9.4% autistic males versus 10% non-autistic males). There was no significant difference in length of stay on PICU between autistic and non-autistic people with psychosis, either by Wilcoxon signed-rank test or adjusted negative binomial regression (median 35 days versus 25 days). There were, however, statistically significant differences in the lifetime lengths of voluntary and involuntary admissions between autistic and non-autistic people (with autistic people having longer durations of admission) (**Figure 1**, **Table 2**).

**Figure 1:**
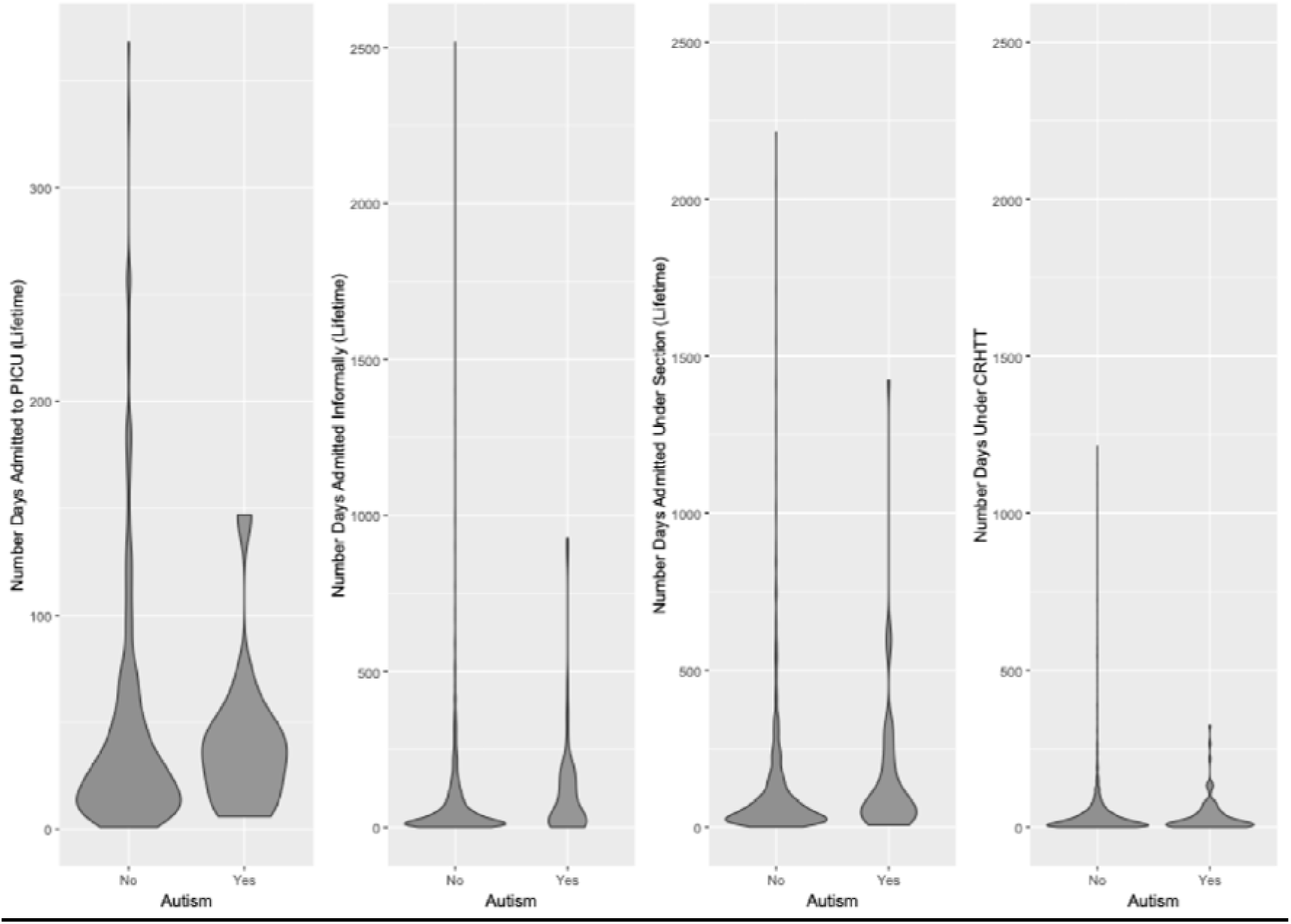
violin plots comparing durations of care for autistic versus non-autistic patients with psychosis in different services. CRHTT=Crisis and Resolution Home Treatment Team.

Although out-of-area admissions were rare, autistic people were more likely to have been admitted out of area across their lifetime (7.4% autistic; 3.5% non-autistic, p<0.05).

### Medication Utilisation

The distribution of lifetime antipsychotic prescribing between autistic and non-autistic people was very similar between groups. These data are presented in **Figure S2 (Supplementary Files)**.

When using logistic regression, autism was not a significant predictor of the number of total lifetime antipsychotics prescribed (median 2 IQR 3 autistic, median 2 IQR 2 non-autistic), use of long-acting injectable use (25.1% autistic, 22.7% non-autistic) or clozapine use (11.6% autistic, 11.7% non-autistic), either in unadjusted or analyses adjusted for age, gender, ethnicity, diagnosis, and socioeconomic status.

### Exploratory Analysis

In our exploratory analyses, we subdivided by learning disability; 26.8% of the autistic group and 1.9% of the non-autistic had a recorded LD diagnosis (**Table S1, see Supplementary Files**). There were 60 people with autism and LD, 164 with autism only, 138 with LD only, and 7047 with neither condition.

In respect to service use outcomes (**Table 5**), the autism-only group had higher use of S136s and the First Response Service in comparison to the other groups. Additionally, the autism-only group used the crisis team at levels comparable to the controls (57.3% autism-only, 61.7% neither), where both the autism-and-LD and LD-only groups had much lower rates of CRHTT usage (10% and 23.2% respectively). In unadjusted regression analyses, autism was associated with the number of EIP contacts, number of CRHTT referrals, S136s, and first response service use; however, in adjusted analyses, autism was no longer an independent predictor of these outcomes. In contrast, LD was associated with the number of EIP contacts, engagement with liaison psychiatry, crisis team use, and first response service use in both unadjusted and adjusted analyses. There were no significant interaction effects.

**Table 3** examines likelihood of hospitalisation following psychosis diagnosis and shows that both the LD-only and autism-and-LD groups had much longer durations in hospital. Autistic individuals’ outcomes more closely resembled those of the group with neither autism or LD, but with much larger variation in lengths of stay (**Table 5)**. However, autism was not a significant predictor of cumulative admission durations, whilst LD was in both the adjusted and unadjusted analyses.

### Sensitivity Analysis

To ensure the validity of our approach, we excluded people awaiting autism diagnosis from our autistic group and re-analysed the outcomes in **Tables 2–3**, as well as medication utilisation (total antipsychotics lifetime, clozapine, and depot utilisation). Overall, the findings were similar to those presented above (see **Supplementary Tables S2-3**).

## Discussion

This study examined data from just over 7,000 patients with psychotic disorders, of whom 3% were autistic or awaiting an autism assessment, and compared outcomes and service use between autistic and non-autistic people. Our study that there are clear differences in the patterns of community and inpatient service use in people who have experienced a first episode of psychosis, even when accounting for covariates.

The prevalence of autism in our sample was perhaps higher than expected, considering national estimates from healthcare data (0.82%) (7). This may reflect the higher risk of psychosis in autistic people, but also could be due to the local early intervention service having some capacity for autism assessments (Kincaid et al., 2017; O’Nions et al., 2023; Treise, Simmons, Marshall, Painter, & Perez, 2021). Under-diagnosis and under-coding may also influence this prevalence, as well as impact on case classification. If autistic people are misclassified, questions about their service use and needs cannot be addressed (and further, the true impact of autism on service use in psychosis may be under-estimated).

These questions on prevalence need to be contextualised in the sample demographics.

The differences in the age composition of the samples (**Table S1)** is notable, possibly reflecting differences in the prevalence and/or diagnosis of autism amongst older adults. There was a higher proportion of males in the autistic group (66.5% male autistic, 49.5% non-autistic), which is in line with traditional gender ratios weighted towards men, but at odds with recent estimates in Swedish data (demonstrating a gender ratio closer to 1M:1F) (Fyfe et al., 2026; Russell et al., 2022). Both of these suggest possible under-diagnosis in older adults and in women.

The finding that autistic people were younger at the age of psychosis diagnosis (even when excluding adults over the age of 60) is perhaps surprising, but could reflect that the supportive infrastructure (clinician contact) that comes with a prior diagnosis. Replication of this analysis in other datasets is required to understand whether this is reproducible or an idiosyncrasy of our local data.

Greater use of crisis telephone services (FRS) and of police MHA Section 136 powers might demonstrate greater unmet need amongst autistic people. However, as seen in the literature on how people from ethnic minority backgrounds with psychosis may enter into the mental health system (higher rates of involuntary admission on first presentation, greater entry to EIS via emergency services/criminal justice), our findings may reflect differences in help-seeking, or in how decompensation manifests (Ghali et al., 2013; Oduola et al., 2019).

This point is particularly relevant when phenomena like autistic burnout and shutdowns are poorly conceptualised in the literature (Mantzalas, Richdale, Li, & Dissanayake, 2024; Phung, Penner, Pirlot, & Welch, 2021).

Furthermore, Section 136 powers are controversial—they are based on one police constable’s opinion and only require someone to appear to be suffering from mental disorder requiring urgent care/control for detention to be legal (UK Government, 1983). There is huge potential for trauma and stigma for adults in crisis coming into contact with police, and poor experiences of the police from autistic people (Borschmann, Gillard, Turner, Chambers, & O’Brien, 2010; Crane, Maras, Hawken, Mulcahy, & Memon, 2016; O’Brien, Sethi, Smith, & Bartlett, 2018). In summary, further understanding on not only precipitants of crisis, but the manifestation of crisis itself and how services respond to autistic people experiencing mental health crisis is required.

Furthermore, while the findings about community services are stark (see **Table 3** unadjusted analyses), they are less so in adjusted regressions (see **Table 5**). Whilst there might be a true lack of effect when considering other factors, there is the possibility of a Type 2 error in these particular adjusted analyses, given that the event numbers are low, the autistic sample is relatively small, combined with data missingness (particularly for ethnicity) to require suppression, and the multiple comparison correction applied.

In line with national data, our findings of higher likelihood and longer total duration of overall inpatient admissions again suggest unmet need (Nisar, Thompson, Boer, Al-Delfi, & Langdon, 2025). This trend is recognised as an unintended consequence of the *Transforming Care* initiative, a drive to move care away from institutions and towards the community, following the 2011 Winterbourne View scandal (a scandal involving mistreatment of people with autism and ID in a care home in England, leading to a government inquiry) (Flynn & Hollins, 2013). The mechanism of this pattern is thought to be the reduction in residential placements with no associated increase in community capacity, thus leading to over-stretched community services which cannot meet the intended aim of admission avoidance for vulnerable people (Nisar et al., 2025). This might help to explain our findings around inpatient admissions (**Table 4**), but also perhaps agree with our exploratory finding that this difference in likelihood of admissions exists despite very similar rates of crisis team referral when looking at autistic and non-autistic people without LD (**Table 5**).

**Table 4:**
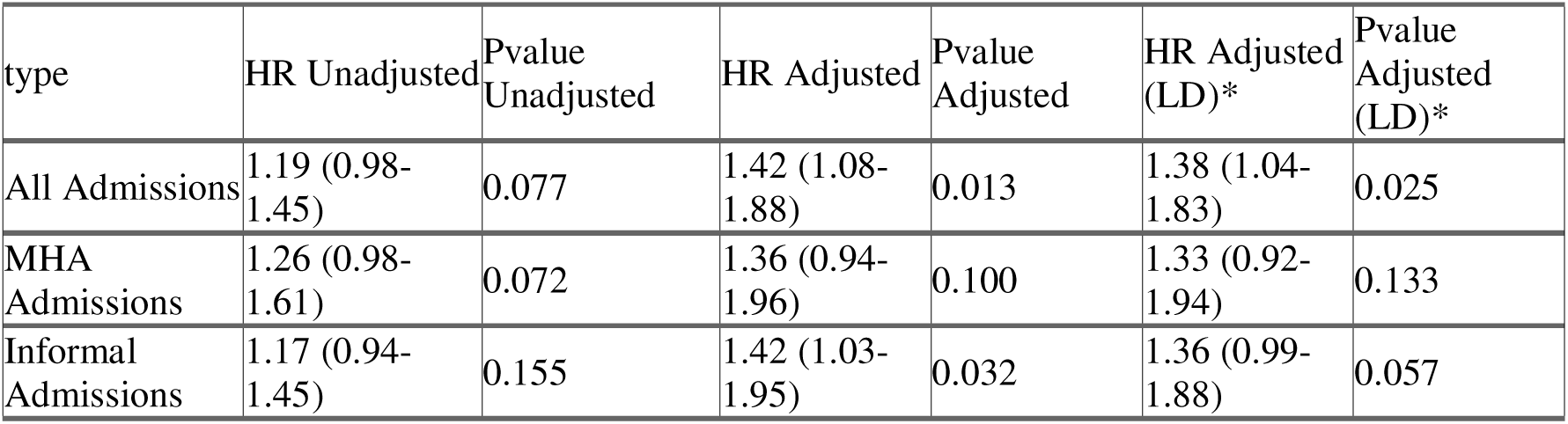
Cox proportional hazard models predicting likelihood of psychiatric hospital admission after diagnosis of a psychotic illness, by autism (adjusted by psychosis diagnostic category, age, ethnicity, gender, IMD decile). HR, hazard ratio. *HR for autism when adjusting by LD.

**Table 5:**
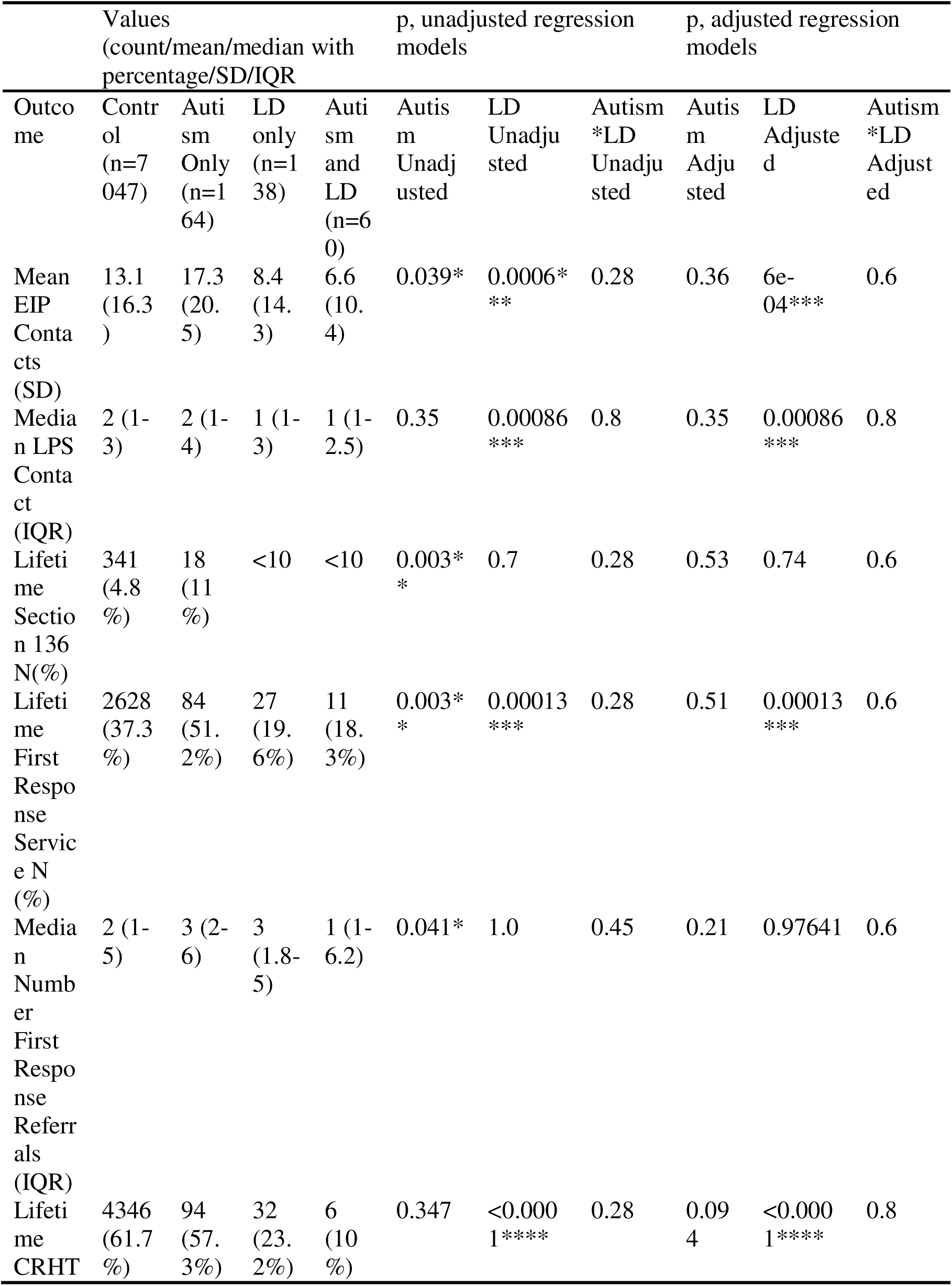
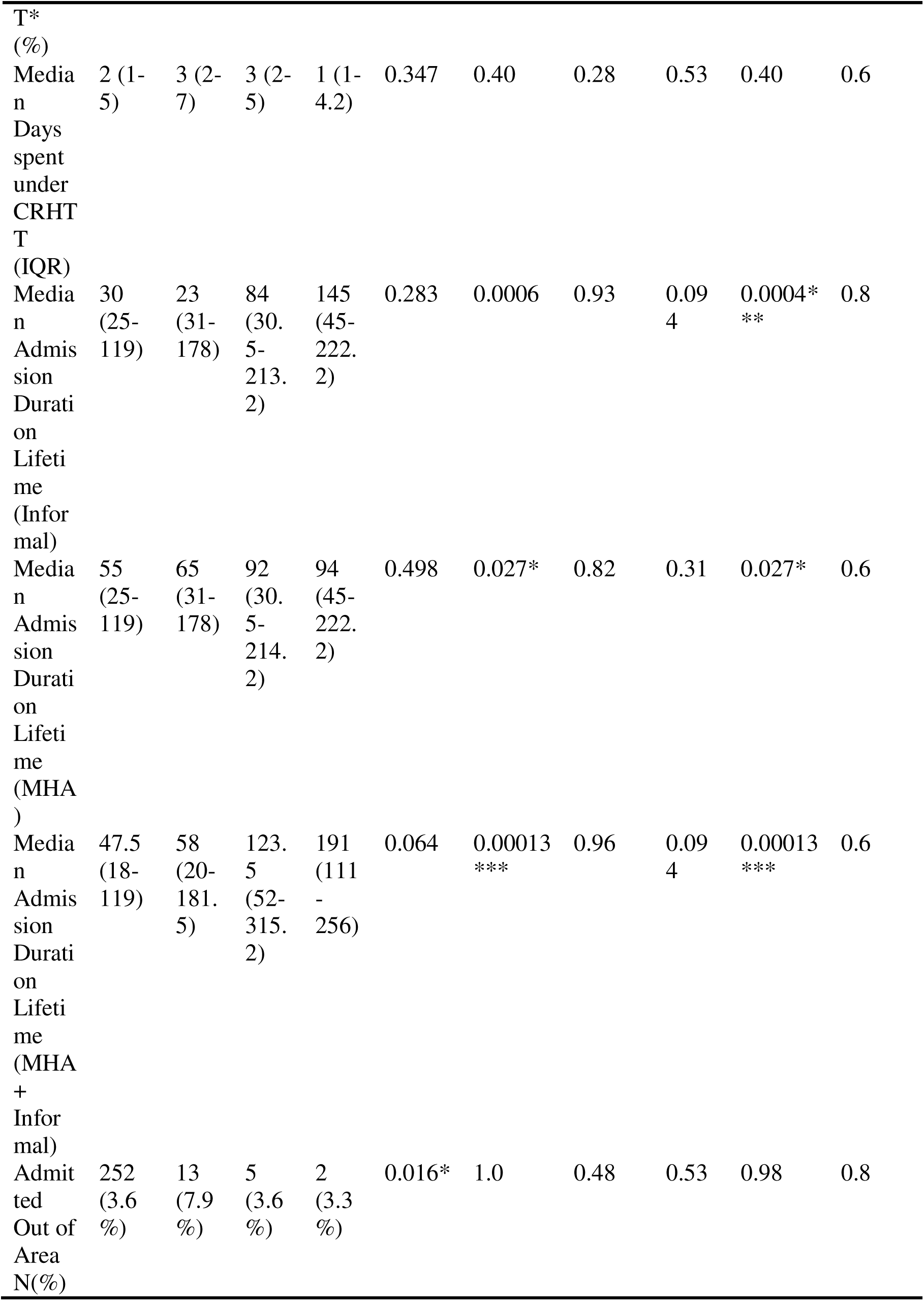
Community and inpatient service use outcomes, subdivided by autism and learning disability (LD), with unadjusted regression model p values (counts=negative binomial, binary=binomial) and adjusted (for age at diagnosis, gender, diagnosis, ethnicity and IMD decile). FDR correction (Benjamini-Hochberg) applied. *CRHTT=Crisis Resolution and Home Treatment Team

Further work to extend this exploratory analysis, with a particular focus on teams whose primary aim is admissions avoidance in these populations (e.g. crisis teams, assertive outreach teams, LD specific teams) is needed to further understand this post Winterbourne View phenomenon.

There are also questions about the greater rates of out-of-area referrals in the autistic-without-LD group compared to other groups, and the small group of autistic people with a longer length of stay on PICUs. It is possible that these may be ‘complex patients’ who cannot be managed in the community following the closure of longer-term placements, and thus appear in other less appropriate settings (e.g. long-term hospitalisation, private locked rehabilitation beds, prisons) (Glasby, Miller, Glasby, Ince, & Konteh, 2024; Glasby, Waring, Miller, Glasby, & Ince, 2025), where subsequent discharge may be difficult (Greenwood, Cooklin, Barbaro, & Miller, 2024; Ince, Glasby, Miller, & Glasby, 2022).

Whilst hospitalisation is sometimes necessary in the treatment of psychosis, our findings suggest that this is more likely for autistic than non-autistic patients (as in the wider literature (McMaughan, Imanpour, Mulcahy, Jones, & Criss, 2023; Tromans, Chester, Kiani, Alexander, & Brugha, 2018)), which is not a desirable outcome for either the patients (inpatient mental health settings often can be particularly traumatic for autistic people) or services who then may struggle to offer safe and therapeutic care (Greenwood et al., 2024; Williams, 2026).

There was a discrepancy in our main and exploratory analyses, in that autistic people were more likely to be admitted following a psychosis diagnosis (**Table 4**) but autism was not a significant predictor of admission length in unadjusted or adjusted regression (**Table 5**).

This may reflect lack of power to detect a small effect, as our sample contained a small number of autistic people who had been admitted (118 autistic people, 3752 non-autistic people), and a large overlap in admission durations (**Table 3**,**5**). Replication of this in a larger dataset would help to understand this further.

Interestingly, learning disability was a strong predictor of both community and inpatient service use, with differences in patterns of use between the autism-LD, autism, and LD groups individually. This may indicate that the influences of autism and LD on outcomes are not simply additive. These findings emphasise the importance of consideration of LD in future work examining the relationship between autism, mental health difficulties, and service interactions.

Given that these results demonstrate very different patterns of service utilisation, it is perhaps curious that there were no measurable differences in prescribing patterns (lifetime number of different antipsychotics prescribed, the use of long-acting injectables or the use of clozapine). This is at odds with an existing finding that autistic adolescents with psychosis were more likely to develop treatment failure (Downs et al., 2017). This discrepancy may relate heterogeneity introduced by a broader age range, and a much broader time period for observation (both of which may affect prescribing practices) (Bosanac & Castle, 2015; Latimer et al., 2014; Nielsen, Dahm, Lublin, & Taylor, 2010).

### Strengths and Limitations

The major strength of this paper is that it leverages clinical data from a large and diverse population, including areas of both affluence and deprivation. Another strength is that this data contains more detail than that available in larger health datasets such as the Mental Health Services Dataset (MHSDS), permitting analysis of specific service utilisation and patterns of admission.

As with all studies based on health records, this study’s major limitation is missingness. It is possible that there were many more cases than those we identified, on account of well-known deficiencies in clinical coding, as well as potential problems of classification bias, particularly amongst the older adults in our sample (Lucyk, Tang, & Quan, 2017). There may also be systematic error in the data, for example particular clinical teams undercoding diagnosis/information.

Another potential weakness is the long time-frame, during which there will have been changes in local policies, clinicians, and attitudes which may impact on clinical practices and thus outcomes. Known examples include (a) the introduction of EIP, which changed the care of people with psychosis over time, and (b) the later introduction of autism screening into local EIP; these would be expected to affect autism diagnosis and referral patterns (Treise et al., 2021), and there may be other such changes. This unmeasured variance may have reduced power.

Future work with greater completeness of ethnicity data in this field is important, given the well-documented impact of ethnicity on interactions with both autism and psychosis services (Eilenberg, Paff, Harrison, & Long, 2019; Fonseca De Freitas et al., 2022; Oduola et al., 2019; Roman-Urrestarazu et al., 2021). In our sample, the large ethnicity missingness and bias towards white patients limits analyses on minority ethnic groups.

### Conclusion

This study characterised outcomes for autistic people following a first episode of psychosis. There were different patterns of service use between autistic and non-autistic people with psychosis, and a notable impact of learning disability. Autistic people required more resources, particularly from emergency and resource-intensive services. Whilst the poor provision of community care for autistic people is recognised and a possible contributor to inpatient admission, it is unclear how to best support autistic people with psychosis and prevent deteriorating mental health, crisis and hospitalisation. Future work should examine and invest in the care of autistic people with psychosis, with established goals of admission avoidance and community care at its core.

## Acknowledgements

we would like to thank our patient/public involvement participants and those clinicians who participated in our engagement activities.

## Funding Statement

Supported by a UK National Institute for Health and Care Research (NIHR) Academic Clinical Fellowship to JHW (ACF-2024-14-015).

EMW was supported by funding from the Autism Centre of Excellence (now Autism Action) in 2023.

RNC’s research is supported by the UK Medical Research Council (MR/Z504816/1). All research at the Department of Psychiatry in the University of Cambridge is supported by the NIHR Cambridge Biomedical Research Centre (NIHR203312) and the NIHR Applied Research Collaboration East of England; the views expressed are those of the author(s) and not necessarily those of the NIHR, the Department of Health and Social Care, or other funders.

## Declarations of interest

RNC receives royalties from Cambridge University Press, Taylor & Francis Group, and Cambridge Enterprise, and consults for Campden Instruments Ltd.

Tamsin Ford’s research group receive funding for research methods consultancy from Place2Be, a third sector organisation providing mental health training and interventions in UK schools. All other authors have nothing to declare.

## Ethics Statement

The CPFT Research Database (CPFTRD) is approved by the UK Research Ethics Service (12/EE/0407, 17/EE/0442, 22/EE/0264) and the project was approved by the CPFTRD Oversight Committee (Z0065).

## Data Availability

Under the terms of NHS ethics approvals, patient-level data is not publicly available. For details of access to the CPFTRD, contact.

## Notes

### Summary of Updates

The abstract in the original version of this manuscript was based on a previous version of the analyses. The abstract has now been updated to reflect the analyses contained within the paper.

## Bibliography

Ahi Üstün, E. S., Yazıcı, S., İlhan, R. S., & Saka, M. C. (2025). Clinical implications of autistic features in the psychosis spectrum: a cross-sectional study using path analysis. BMC Psychiatry 2025 25:1, 25(1), 1–15. doi:10.1186/S12888-025-07377-Z

Ajnakina, O., Stubbs, B., Francis, E., Gaughran, F., David, A. S., Murray, R. M., & Lally, J. (2020). Hospitalisation and length of hospital stay following first-episode psychosis: systematic review and meta-analysis of longitudinal studies. Psychological Medicine, 50(6), 991–1001. doi:10.1017/S0033291719000904

Borschmann, R. D., Gillard, S., Turner, K., Chambers, M., & O’Brien, A. (2010). Section 136 of the Mental Health Act: A new literature review. *Medicine*, Science and the Law, 50(1), 34–39. doi:10.1258/MSL.2009.009004;SUBPAGE:STRING:FULL

Bosanac, P., & Castle, D. J. (2015). Why are long-acting injectable antipsychotics still underused? BJPsych Advances, 21(2), 98–105. doi:10.1192/apt.bp.114.013565

Cardinal, R. N. (2017). Clinical records anonymisation and text extraction (CRATE): an open-source software system. BMC Medical Informatics and Decision Making, 17(1). doi:10.1186/S12911-017-0437-1

Cardinal, R. N., Savulich, G., Mann, L. M., & Fernández-Egea, E. (2015). Association between antipsychotic/antidepressant drug treatments and hospital admissions in schizophrenia assessed using a mental health case register. Npj Schizophrenia 2015 1:1, 1(1), 15035-. doi:10.1038/npjschz.2015.35

Chisholm, K., Pelton, M., Duncan, N., Kidd, K., Wardenaar, K. J., Upthegrove, R., … Wood, S. J. (2019). A cross-sectional examination of the clinical significance of autistic traits in individuals experiencing a first episode of psychosis. Psychiatry Research, 282, 112623. doi:10.1016/J.PSYCHRES.2019.112623

Correll, C. U., Solmi, M., Croatto, G., Schneider, L. K., Rohani-Montez, S. C., Fairley, L., … Tiihonen, J. (2022). Mortality in people with schizophrenia: a systematic review and meta-analysis of relative risk and aggravating or attenuating factors. World PsychiatrylJ: Official Journal of the World Psychiatric Association (WPA), 21(2), 248–271. doi:10.1002/WPS.20994

Crane, L., Maras, K. L., Hawken, T., Mulcahy, S., & Memon, A. (2016). Experiences of Autism Spectrum Disorder and Policing in England and Wales: Surveying Police and the Autism Community. Journal of Autism and Developmental Disorders 2016 46:6, 46(6), 2028–2041. doi:10.1007/s10803-016-2729-1

de Pablo, G. S., Guinart, D., Armendariz, A., Aymerich, C., Catalan, A., Alameda, L., … Correll, C. U. (2024). Duration of Untreated Psychosis and Outcomes in First-Episode Psychosis: Systematic Review and Meta-analysis of Early Detection and Intervention Strategies. Schizophrenia Bulletin, 50(4), 771–783. doi:10.1093/SCHBUL/SBAE017

de Winter, L., Couwenbergh, C., van Weeghel, J., Hasson-Ohayon, I., Vermeulen, J. M., Mulder, C. L., … Veling, W. (2022). Changes in social functioning over the course of psychotic disorders–A meta-analysis. Schizophrenia Research, 239, 55–82. doi:10.1016/j.schres.2021.11.010

de Winter, L., Vermeulen, J. M., Couwenbergh, C., van Weeghel, J., Hasson-Ohayon, I., Mulder, C. L., … de Haan, L. (2023). Short- and long-term changes in symptom dimensions among patients with schizophrenia spectrum disorders and different durations of illness: A meta-analysis. Journal of Psychiatric Research, 164, 416–439. doi:10.1016/j.jpsychires.2023.06.031

Downs, J. M., Lechler, S., Dean, H., Sears, N., Patel, R., Shetty, H., … Pina-Camacho, L. (2017). The association between comorbid autism spectrum disorders and antipsychotic treatment failure in early-onset psychosis: A historical cohort study using electronic health records. Journal of Clinical Psychiatry, 78(9), e1233–e1241. doi:10.4088/JCP.16M11422

Eilenberg, J. S., Paff, M., Harrison, A. J., & Long, K. A. (2019, May 1). Disparities Based on Race, Ethnicity, and Socioeconomic Status Over the Transition to Adulthood Among Adolescents and Young Adults on the Autism Spectrum: a Systematic Review. Current Psychiatry Reports. Current Medicine Group LLC 1. doi:10.1007/s11920-019-1016-1

Fern, M., Regan, C., & Lucas-Motley, H. (2025). Quality Standards for Early Intervention in Psychosis Services (Third Edition). Retrieved from www.rcpsych.ac.uk/EIPN

Flynn, M., & Hollins, S. (2013). Acting on the lessons of Winterbourne View Hospital. BMJ, 346(7890). doi:10.1136/BMJ.F18

Fonseca De Freitas, D., Pritchard, M., Shetty, H., Khondoker, M., Nazroo, J., Hayes, R. D., & Bhui, K. (2022). Ethnic inequities in multimorbidity among people with psychosis: a retrospective cohort study. Epidemiology and Psychiatric Sciences, 31. doi:10.1017/S2045796022000385

Fyfe, C., Winell, H., Dougherty, J., Gutmann, D. H., Kolevzon, A., Marrus, N., … Sandin, S. (2026). Time trends in the male to female ratio for autism incidence: population based, prospectively collected, birth cohort study. BMJ, 392, e084164. doi:10.1136/BMJ-2025-084164

Ghali, S., Fisher, H. L., Joyce, J., Major, B., Hobbs, L., Soni, S., … Johnson, S. (2013). Ethnic variations in pathways into early intervention services for psychosis. The British Journal of Psychiatry, 202(4), 277–283. doi:10.1192/bjp.bp.111.097865

Glasby, J., Miller, R., Glasby, A. M., Ince, R., & Konteh, F. (2024). ‘Why are we stuck in hospital?’ Barriers to people with learning disabilities/autistic people leaving ‘long-stay’ hospital: a mixed methods study. Health and Social Care Delivery Research, 12(3), 1–119. doi:10.3310/HBSH7124

Glasby, J., Waring, J., Miller, R., Glasby, A. M., & Ince, R. (2025). Out of Sight, Out of Mind—Explaining and Challenging the ReLJInstitutionalisation of People With Learning Disabilities and/or Autistic People. Sociology of Health & Illness, 47(2), e70009. doi:10.1111/1467-9566.70009

Greenwood, E., Cooklin, A., Barbaro, J., & Miller, C. (2024). Autistic patients’ experiences of the hospital setting: A scoping review. Journal of Advanced Nursing, 80(3), 908–923. doi:10.1111/jan.15880

Ince, R., Glasby, J., Miller, R., & Glasby, A. M. (2022). ‘Why are we stuck in hospital?’ Understanding delayed hospital discharges for people with learning disabilities and/or autistic people in long-stay hospitals in the UK. Health and Social Care in the Community, 30(6), e3477–e3492. doi:10.1111/hsc.13964

Jeong, J. H., Kim, S. W., Yu, J. C., Won, S. H., Lee, S. H., Kim, S. H., … Lee, K. Y. (2024). Clinical, cognitive, and functional characteristics of recent-onset psychosis with autistic features: A 2-year longitudinal study. Schizophrenia Research, 270, 304–316. doi:10.1016/j.schres.2024.06.009

Kincaid, D. L., Doris, M., Shannon, C., & Mulholland, C. (2017). What is the prevalence of autism spectrum disorder and ASD traits in psychosis? A systematic review. Psychiatry Research, 250, 99–105. doi:10.1016/j.psychres.2017.01.017

Latimer, E. A., Naidu, A., Moodie, E. E. M., Clark, R. E., Malla, A. K., Tamblyn, R., & Wynant, W. (2014). Variation in long-term antipsychotic polypharmacy and high-dose prescribing across physicians and hospitals. Psychiatric Services, 65(10), 1210–1217. doi:10.1176/appi.ps.201300217

Lucyk, K., Tang, K., & Quan, H. (2017). Barriers to data quality resulting from the process of coding health information to administrative data: a qualitative study. BMC Health Services Research 2017 17:1, 17(1), 766-. doi:10.1186/S12913-017-2697-Y

Mantzalas, J., Richdale, A. L., Li, X., & Dissanayake, C. (2024). Measuring and validating autistic burnout. Autism Research, 17(7), 1417–1449. doi:10.1002/aur.3129

Martini, M. I., Kuja-Halkola, R., Butwicka, A., Du Rietz, E., D’Onofrio, B. M., Happé, F., … Taylor, M. J. (2022). Sex Differences in Mental Health Problems and Psychiatric Hospitalization in Autistic Young Adults. JAMA Psychiatry, 79(12), 1188–1198. doi:10.1001/JAMAPSYCHIATRY.2022.3475

McMaughan, D. J., Imanpour, S., Mulcahy, A., Jones, J., & Criss, M. M. (2023). Mental health-related hospitalizations among adolescents and emerging adults with autism in the United States: A retrospective, cross-sectional analysis of national hospital discharge data. AutismlJ: The International Journal of Research and Practice, 27(6), 1702–1715. doi:10.1177/13623613221143592

Moreno, C., Nuevo, R., Chatterji, S., Verdes, E., Arango, C., & Ayuso-Mateos, J. L. (2013). Psychotic symptoms are associated with physical health problems independently of a mental disorder diagnosis: results from the WHO World Health Survey. World Psychiatry, 12(3), 251. doi:10.1002/WPS.20070

Morris, S., Hill, S., Brugha, T., & McManus, S. (2025). Adult Psychiatric Morbidity Survey: Survey of Mental Health and Wellbeing, England, 2023/4 - NHS England Digital. Retrieved from https://digital.nhs.uk/data-and-information/publications/statistical/adult-psychiatric-morbidity-survey/survey-of-mental-health-and-wellbeing-england-2023-24

National Institute for Health and Care Excellence. (2015). Quality statement 1: Referral to early intervention in psychosis services | Psychosis and schizophrenia in adults | Quality standards | NICE. Retrieved 5 November 2025, from https://www.nice.org.uk/guidance/qs80/chapter/quality-statement-1-referral-to-early-intervention-in-psychosis-services

NHS Digital. (2025). Autism Statistics, October 2024 to September 2025. Retrieved from https://digital.nhs.uk/data-and-information/publications/statistical/autism-statistics/october-2024-to-september-2025

Nielsen, J., Dahm, M., Lublin, H., & Taylor, D. (2010). Psychiatrists’ attitude towards and knowledge of clozapine treatment. *Journal of Psychopharmacology (Oxford*, England), 24(7), 965–971. doi:10.1177/0269881108100320

Nikolić, N., Sculthorpe, C., Stock, J., Stevens, D., & Eccles, J. (2025). Determining unmet need: clinical relevance of suspected neurodivergence in first-episode psychosis. BJPsych Bulletin, 49(6), 385–390. doi:10.1192/bjb.2024.64

Nisar, A., Thompson, P. A., Boer, H., Al-Delfi, H., & Langdon, P. E. (2025). Factors Affecting Psychiatric Bed Utilisation by People With Intellectual Disabilities: A Time Series Analysis Using the English National Mental Health Services Data Set. Journal of Intellectual Disability Research, 69(11), 1251. doi:10.1111/JIR.70003

O’Brien, A., Sethi, F., Smith, M., & Bartlett, A. (2018). Public mental health crisis management and Section 136 of the Mental Health Act. Journal of Medical Ethics, 44(5), 349–353. doi:10.1136/MEDETHICS-2016-103994

Oduola, S., Craig, T. K. J., Das-Munshi, J., Bourque, F., Gayer-Anderson, C., & Morgan, C. (2019). Compulsory admission at first presentation to services for psychosis: does ethnicity still matter? Findings from two population-based studies of first episode psychosis. Social Psychiatry and Psychiatric Epidemiology, 54(7), 871–881. doi:10.1007/s00127-019-01685-y

Oduola, S., Craig, T. K. J., Iacoponi, E., Macdonald, A., & Morgan, C. (2024). Sociodemographic and clinical predictors of delay to and length of stay with early intervention for psychosis service: findings from the CRIS-FEP study. Social Psychiatry and Psychiatric Epidemiology, 59(1), 25–36. doi:10.1007/S00127-023-02522-Z

Office for National Statistics. (2022, November 29). Ethnic group, England and Wales: Census 2021. Retrieved 3 February 2026, from https://www.ons.gov.uk/peoplepopulationandcommunity/culturalidentity/ethnicity/bulletins/ethnicgroupenglandandwales/census2021

O’Nions, E., Lewer, D., Petersen, I., Brown, J., Buckman, J. E. J., Charlton, R., … Stott, J. (2024). Estimating life expectancy and years of life lost for autistic people in the UK: a matched cohort study. The Lancet Regional Health - Europe, 36, 100776. doi:10.1016/j.lanepe.2023.100776

O’Nions, E., Petersen, I., Buckman, J. E. J., Charlton, R., Cooper, C., Corbett, A., … Stott, J. (2023). Autism in England: assessing underdiagnosis in a population-based cohort study of prospectively collected primary care data. The Lancet Regional Health - Europe, 29. doi:10.1016/j.lanepe.2023.100626

Peritogiannis, V., Gogou, A., & Samakouri, M. (2020). Very long-term outcome of psychotic disorders. International Journal of Social Psychiatry, 66(7), 633–641. doi:10.1177/0020764020922276;CTYPE:STRING:JOURNAL

Phung, J., Penner, M., Pirlot, C., & Welch, C. (2021). What I Wish You Knew: Insights on Burnout, Inertia, Meltdown, and Shutdown From Autistic Youth. Frontiers in Psychology, 12, 741421. doi:10.3389/fpsyg.2021.741421

Puntis, S., Minichino, A., De Crescenzo, F., Harrison, R., Cipriani, A., & Lennox, B. (2020). Specialised early intervention teams (extended time) for recent-onset psychosis. Cochrane Database of Systematic Reviews, 2020(11). doi:10.1002/14651858.CD013287.PUB2/MEDIA/CDSR/CD013287/URN:X-WILEY:14651858:MEDIA:CD013287:CD013287-CMP-001.25

Radev, S., Freeth, M., & Thompson, A. R. (2024). How healthcare systems are experienced by autistic adults in the United Kingdom: A meta-ethnography. Autism, 28(9), 2166–2178. doi:10.1177/13623613241235531

Roman-Urrestarazu, A., Van Kessel, R., Allison, C., Matthews, F. E., Brayne, C., & Baron-Cohen, S. (2021). Association of Race/Ethnicity and Social Disadvantage With Autism Prevalence in 7 Million School Children in England. JAMA Pediatrics, 175(6). doi:10.1001/JAMAPEDIATRICS.2021.0054

Russell, G., Stapley, S., Newlove-Delgado, T., Salmon, A., White, R., Warren, F., … Ford, T. (2022). Time trends in autism diagnosis over 20 years: a UK population-based cohort study. Journal of Child Psychology and Psychiatry and Allied Disciplines, 63(6), 674–682. doi:10.1111/jcpp.13505

Treise, C., Simmons, C., Marshall, N., Painter, M., & Perez, J. (2021). Autism Spectrum Disorder in Early Intervention in Psychosis Services: Implementation and Findings of a 3-step Screening and Diagnostic Protocol. Journal of Psychiatric Practice, 27(1), 23–32. doi:10.1097/PRA.0000000000000525

Tromans, S., Chester, V., Kiani, R., Alexander, R., & Brugha, T. (2018). The Prevalence of Autism Spectrum Disorders in Adult Psychiatric Inpatients: A Systematic Review. Clinical Practice and Epidemiology in Mental Health, 14(1), 177–187. doi:10.2174/1745017901814010177

UK Government. Mental Health Act (1983) (1983). Statute Law Database.

Vaquerizo-Serrano, J., Salazar de Pablo, G., Singh, J., & Santosh, P. (2022). Autism Spectrum Disorder and Clinical High Risk for Psychosis: A Systematic Review and Meta-analysis. Journal of Autism and Developmental Disorders, 52(4), 1568–1586. doi:10.1007/S10803-021-05046-0

Vogan, V., Lake, J. K., Tint, A., Weiss, J. A., & Lunsky, Y. (2017). Tracking health care service use and the experiences of adults with autism spectrum disorder without intellectual disability: A longitudinal study of service rates, barriers and satisfaction. Disability and Health Journal, 10(2), 264–270. doi:10.1016/j.dhjo.2016.11.002

Ward, John H., Weir, E., Allison, C., & Baron-Cohen, S. (2023). Increased rates of chronic physical health conditions across all organ systems in autistic adolescents and adults. Molecular Autism, 14(1), 1–20. doi:10.1186/S13229-023-00565-2/FIGURES/1

Ward, John Headley, Lewis, J. R., Weir, E. M., Ford, T. J., & Cardinal, R. N. (2026). Comparing outcomes following a first episode of psychosis in autistic and non-autistic people: a clinical retrospective cohort study. MedRxiv, 2026.06.01.26354576. doi:10.64898/2026.06.01.26354576

Weir, E., Allison, C., Warrier, V., & Baron-Cohen, S. (2021). Increased prevalence of non-communicable physical health conditions among autistic adults. Autism, 25(3), 681–694. doi:10.1177/1362361320953652

White, C., Stirling, J., Hopkins, R., Morris, J., Montague, L., Tantam, D., & Lewis, S. (2009). Predictors of 10-year outcome of first-episode psychosis. Psychological Medicine, 39(9), 1447–1456. doi:10.1017/S003329170800514X

Williams, R. (2026). ‘It’s not fair, this isn’t what an autistic person should go through’: Experiences of autistic adults on inpatient mental health wards. Autism. doi:10.1177/13623613251412722

Zheng, S., Chua, Y. C., Tang, C., Tan, G. M. Y., Abdin, E., Lim, V. W. Q., … Magiati, I. (2021). Autistic traits in first-episode psychosis: Rates and association with 1-year recovery outcomes. Early Intervention in Psychiatry, 15(4), 849–855. doi:10.1111/EIP.13021

Zheng, Z., Zheng, P., & Zou, X. (2018). Association between schizophrenia and autism spectrum disorder: A systematic review and meta-analysis. Autism Research, 11(8), 1110–1119. doi:10.1002/AUR.1977

Zumstein, N., & Riese, F. (2020). Defining Severe and Persistent Mental Illness—A Pragmatic Utility Concept Analysis. Frontiers in Psychiatry, 11, 648. doi:10.3389/FPSYT.2020.00648

