## Supplementary Files for "Comparing outcomes following a first episode of psychosis in autistic and non-autistic people: a clinical retrospective cohort study"

**Supplementary Materials for Paper**

**Supplementary Methods:**

**Community Service Use Data Extraction**

Information on service used was derived differently depending on the type of service accessed and the origin of the clinical record (RiO or SystmOne). Referrals to clinical teams, such as EIP (locally known as ‘Cambridgeshire and Peterborough Assessing Managing and Enhancing Outcomes [EIP]’), or Liaison Psychiatry, were identified via referral records, with data on appointments also extracted (e.g. service, attendance stautus).

Data on urgent mental health support phone line use (accessed via the national NHS 111 crisis line, known locally as the ‘First Response Service’ [FRS]) was extracted from completed forms in health records. These were de-duplicated by date/time.

Uses of Section 136 (S136) of the Mental Health Act (MHA) were also recorded. Section 136 powers allow the police to remove someone with a suspected mental health disorder needing urgent help from a public place to a place of safety (e.g. an acute hospital emergency department or a mental health dedicated ‘S136 Suite’) (UK Government, 1983). The dates of each S136 event per person were extracted.

Uses of the Crisis and Resolution Home Treatment Team (CRHTT) were identified through referrals, with the length of care received calculated based on referral and discharge dates.

**Inpatient Service Use Data Extraction**

Data on inpatient admissions were extracted for all patients, including the dates of admissions and MHA usage. Uses of the MHA (‘sections’) denote an involuntary/detained admission, where other admissions are voluntary or ‘informal’. In our analyses of MHA frameworks, we only included sections which could be used to detain a patient for at least 28 days (i.e. civil sections [s2/s3]; forensic sections [s35/s37/s45A/s47]). For context, the major MHA provisions used to treat patients in the UK are Section 2 (permitting detention for up to 28 days for assessment and treatment, given sufficient risk) and Section 3 (permitting detention for up to 6 months for treatment, if certain other criteria are also met).

**Out-of-Area Admissions**

An ‘out-of-area admission’ denotes someone being admitted outside of the CPFT estate, often in a subcontracted private facility. Reasons include local bed availability or a service required that is not offered by CPFT (e.g. female psychiatric intensive care beds). Data on out-of-area admissions were extracted in two ways. Firstly, data were extracted from SystmOne via the dedicated ‘Out of Area Placements’ table. Given no such equivalent table existed in the RiO records, people who had been placed out of area were identified by clinical document titles. Document titles of interest were documents containing names of known private providers used by the mental health trust (confirmed with a relevant manager)

List of out-of-area providers:

- Audley, Benfleet, Broomhill, Cygnet, Elysium, Frinton, Heygate, Magna House, OOA PICU, Priory, Southern Hill.

**List of antipsychotics:**

- Amisulpiride, aripiprazole, asenapine, chlorpromazine, chlorprothixene, clozapine, droperidol, flupentixol, fluphenazine, haloperidol, iloperidone, levomepromazine, lurasidone, methotrimeprazine, molindone, olanzapine, paliperidone, pericyazine, perphenazine, pimozide, promazine, quetiapine, risperidone, sertindole, sulpiride, thioridazine, thiothixene, trifluoperazine, ziprasidone, zotepine, zuclopenthixol

**Identifying Long-Acting Injectables (LAIs)**

Long-acting injectable (LAI, or depot) antipsychotic prescriptions were extracted firstly from a list of known LAIs and then on three further criteria. These were: if the drug name extract from the NLP included ‘depot’ in the title (e.g. haloperidol depot), if the drug name extract included a known injectable form (e.g. haloperidol decanoate, Risperdal Consta), or if the dose of the drug was above that of a typical daily oral dose (e.g. haloperidol doses above 20mg).

**Supplementary Results:**

**Table S1: Year/age of diagnosis of a psychotic disorder, and presence of intellectual (learning) disability, by group**

| Variable | Non-autistic (N=7185) | Autistic (N=224) |
| --- | --- | --- |
| 0–17 | 84 (1.2%) | 12 (5.4%) |
| 18–29 | 1322 (18.4%) | 89 (39.7%) |
| 30–39 | 1396 (19.4%) | 51 (22.8%) |
| 40–49 | 1216 (16.9%) | 35 (15.6%) |
| 50–59 | 1074 (14.9%) | 26 (11.6%) |
| 60+ | 2088 (29.1%) | 11 (4.9%) |
| Mild/other/unspecified learning disability | 89(1.2%) | 31(12.9%) |
| Moderate learning disability | 36(0.5%) | 16(7.1%) |
| Severe learning disability | 13(0.2%) | 13(5.8%) |
| 1950-2019 | 5688 (79.2%) | 179 (79.9%) |
| 2020-26 | 1492 (20.8%) | 45 (20.1%) |

**Table S2: Table 1 with people awaiting autism diagnoses excluded.**

| **Variable** | **Non-autistic (N=7185)** | **Autistic (N=185)** | **P.value** |
| --- | --- | --- | --- |
| Acute and transient pscyhotic disorders (including other nonorganic psychotic disorders) | 481 (6.7%) | 14 (7.6%) | 0.11 |
| Bipolar affective disorder (including mania) (including mania) | 2340 (32.6%) | 69 (37.3%) | 1 |
| Mental & behavioral disorders due to psychoactive substance use | 184 (2.6%) | <10 (<5%) | 0.48 |
| Persistent delusional disorders | 322 (4.5%) | <10 (<5%) | 0.54 |
| Psychotic depression | 884 (12.3%) | 13 (7%) | 0.04* |
| Schizoaffective disorders | 445 (6.2%) | 14 (7.6%) | 0.33 |
| Schizophrenia | 1696 (23.6%) | 39 (21.1%) | 0.3 |
| Schizotypal disorder | 36 (0.5%) | <10 (<5%) | 0.2 |
| Unspecified nonorganic psychosis | 797 (11.1%) | 28 (15.1%) | 0.75 |
| Mean Age at Diagnosis (SD) | 48 (19) | 34.1 (13.5) | p<0.0001* |
| Male N (%) | 3553 (49.5%) | 124 (67%) |  |
| Asian | 356 (5%) | <10 (<5%) |  |
| Black | 175 (2.4%) | <10 (<5%) |  |
| Missing | 1702 (23.7%) | 22 (11.9%) |  |
| Mixed | 122 (1.7%) | <10 (<5%) |  |
| Other | 783 (10.9%) | 16 (8.6%) |  |
| White | 4047 (56.3%) | 133 (71.9%) |  |
| Mean IMD (SD) | 5.7 (2.7) | 5.7 (2.6) | 0.82 |
| N Died (%) | 833 (11.6%) | <10 (<5%) | p<0.0001* |
| Mean Age at death | 70.9 | 49.2 | 0.09 |

**Table S3: Table 2 with people awaiting autism diagnoses excluded.**

| **Variable** | **Non-autistic (N=7185)** | **Autistic (N=185)** | **P.Value** |
| --- | --- | --- | --- |
| Variable | Non-autistic (N=7185) | Autistic (N=248) | P.Value |
| Referred to EIP (%) | 1268 (17.6%) | 38 (20.5%) | 0.31 |
| Inappropriate Referral (%) | 198 (15.6%) | 8 (21.1%) | 0.36 |
| Treatment Completed (%) | 565(44.6%) | 22(57.9%) | 0.1 |
| Mean Contact (SD) | 13 (16.3) | 14.9 (19.8) | 0.28 |
| Seen by LPS (%) | 3418 (47.6%) | 70 (37.8%) | 0.01* |
| Median LPS Contact (IQR) | 2 (2) | 2 (2) | 0.81 |
| Lifetime S136 (%) | 349 (4.9%) | 15 (8.1%) | 0.04* |
| S136 Count Median (IQR) | 2 (3) | 3 (4.5) | 0.25 |
| Lifetime FRS (%) | 2655 (37%) | 70 (37.8%) | 0.81 |
| FRS Count Median (IQR) | 2 (15) | 3 (19.8) | 0.15 |
| Lifetime CRHTT (%) | 4378 (60.9%) | 79 (42.7%) | p<0.0001 |
| Admitted to PICU N Males(%) | 354 (10%) | 11 (8.9%) | 0.69 |
| Median PICU LoS (IQR) | 25 (37) | 29 (28) | 0.92 |
| Admitted Informally N(%) | 2760 (38.4%) | 80 (43.2%) | 0.18 |
| Median Admission Duration (Informal) | 31 (69) | 54.5 (135.75) | 0.01* |
| Admitted Under MHA N(%) | 1865 (26%) | 59 (31.9%) | 0.07 |
| Median Admission Duration (MHA) | 55 (97) | 69 (98.5) | 0.06 |
| Admitted Ever (%) | 3752 (52.2%) | 102 (55.1%) | 0.43 |
| Median Admission Duration (Total) | 49 (102) | 114 (177.75) | p<0.0001 |
| Admitted Out of Area N(%) | 257 (3.6%) | 13 (7%) | 0.01* |

**Figure S1:** timing of autism diagnosis in relation to psychosis diagnosis

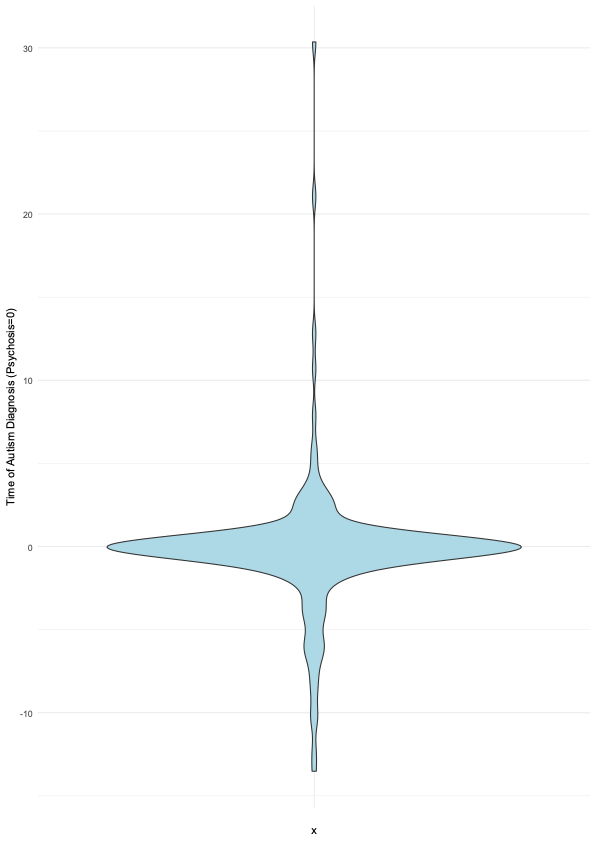

**Figure S2:** bar plot demonstrating the number of different antipsychotics prescribed in autistic and non-autistic people with psychosis.

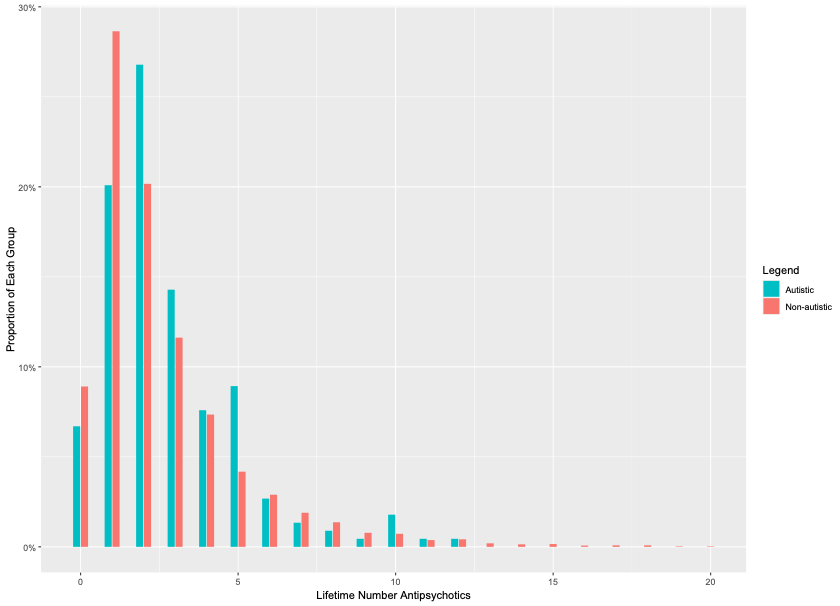
